# Evaluation of *indirect* damage and damage saturation effects in dose-response curves of hypofractionated radiotherapy of early-stage NSCLC and brain metastases

**DOI:** 10.1101/2020.08.26.20182287

**Authors:** Araceli Gago-Arias, Sara Neira, Miguel Pombar, Antonio Gómez-Caamaño, Juan Pardo-Montero

**Affiliations:** Group of Medical Physics and Biomathematics, Instituto de Investigación Sanitaria de Santiago (IDIS), Santiago de Compostela, Spain; Department of Medical Physics, Complexo Hospitalario Universitario de Santiago de Compostela, Spain; Institute of Physics, Pontificia Universidad Católica, Chile; Group of Molecular Imaging and Oncology, Instituto de Investigación Sanitaria de Santiago (IDIS), Santiago de Compostela, Spain; Department of Radiotherapy, Complexo Hospitalario Universitario de Santiago de Compostela, Spain

**Keywords:** SBRT, radiobiological modeling, vascular damage, NSCLC, brain metastases

## Abstract

**Background:** In this work we aim to investigate the possible contribution of *indirect* damage and damage saturation to tumor control probabilities (TCP) obtained with SBRT/SRS treatments for early-stage NSCLC and brain metastases.

**Methods:** We have constructed a dataset of early-stage NSCLC and brain metastases response to different fractionations. Dose-response curves were fitted to models based on the linear-quadratic (LQ), the linear-quadratic-linear (LQL), and phenomenological modifications of the LQ model to account for indirect cell damage. We used the Akaike-Information-Criterion formalism to compare performance, and studied the stability of the results with changes in fitting parameters and perturbations on dose/TCP values.

**Results:** In NSCLC, a modified LQ model with a beta-term increasing with dose yields better results than the LQ model. This rank remains consistent when different fitting parameters are changed, and only the inclusion of very fast accelerated proliferation can eliminate the superiority of the modified LQ. In brain, the LQL model yields the best-fits, and the ranking is not affected by variations of fitting parameters or dose/TCP perturbations.

**Conclusions:** A modification of the LQ model with a beta-term increasing with dose provides better fits to NSCLC dose-response curves. For brain metastases, the LQL provides the best fit. This may be interpreted as a net contribution of *indirect* damage in NSCLC, and damage saturation in brain metastases. The results for NSCLC are borderline significant, while those for brain are clearly significant. Our results can assist on the design of optimal radiotherapy for NSCLC and brain metastases, aiming at avoiding over/under-treatment. Dose prescription to such tumors may be reevaluated according to the reported evidence.

## Background

Improvements in the accuracy of radiotherapy delivery systems have lead to a situation where highdose fractions can be safely delivered. This has driven the implementation of Stereotactic Body RadioTherapy (SBRT) and Stereotactic Radiation Surgery (SRS) **[1-3]**, hypofractionated radiotherapy techniques where doses per fraction can reach up to 35-40 Gy, much higher than the typical 2 Gy per fraction delivered in conventional radiotherapy.

For the past thirty years, the linear-quadratic (LQ) model **[4]** has guided the analysis of radiotherapy outcomes and the design of novel fractionations. However, motivated by the increasing use of hypofractionation, there is an intense debate on the radiobiology of large radiation doses and the applicability of the LQ model **[5-9]**. Several experiments have shown that high-doses per fraction lose effectiveness when compared to expected values from low-dose extrapolations, which may be due to higher effective repair rates **[10]**. Variations of the LQ model were developed to describe this behavior **[10-15]**, which we will refer to as *“damage saturation”*. However, recent experimental results also show that high doses per fraction may trigger new mechanisms of cell death, sometimes called *indirect* cell death. *Indirect* cell death seems to be caused by two different mechanisms, induction of vascular damage that can lead to cell starvation and death **[16-19]**, and the activation of an immune-response against the tumor **[20-26]**.

Such effects seem to be well established experimentally, but there is still some controversy on their contribution to tumor control rates. For example, recent pre-clinical studies show that endothelial cell death may not play a significant role in tumor control in experiments with genetically modified mice with radiation-sensitive endothelial cells **[27, 28]**. On the other hand, on the clinical side, there seems to be little evidence of indirect cell damage contributing to tumor control obtained with SBRT/SRS treatments. An analysis by Brown and colleagues of outcomes of SBRT concluded that results could be explained by dose escalation alone **[29]**. Shuryak and colleagues also analyzed outcomes of SBRT (lung cancer) and SRS (brain metastases), concluding that the LQ model provides better fits to data than modifications of the LQ model with decreasing effectiveness with increasing dose (like the LQL and the USC models) **[30]**. However, they did not investigate the possibility of an increase of effectiveness with dose-per-fraction, as would be caused by indirect cell damage.

In this work we further explore the possible contribution of indirect cell death effects and/or damage saturation to tumor control probabilities obtained with SBRT/SRS treatments of Non Small Cell Lung Cancer (NSCLC) and brain metastases. We have constructed a large dataset of treatment outcomes to different fractionation schemes, building on the analyses reported in references **[30, 31]**, and including more studies/fractionations. The methodology that we follow consists on fitting biomathematical tumor control probabilities (TCP) models, typically constructed from the LQ-model, to experimental TCP curves. We hypothesize that the effect of indirect cell death can be described by an *ad hoc* modification of the LQ-model with the following general form:

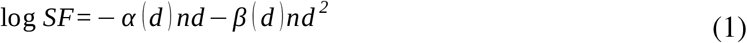

In this expression α*(d*) and *β(d*) represent a dependence of the *α-β* parameters on the dose per fraction, d. For example, if indirect damage is more important for large doses, *α(d*) and *β(d*) should have the form of monotonically-increasing functions.

We explore the goodness of fits obtained with simple *ad hoc* variations of the LQ-model along the line reported in equation (1), and compare them with results obtained with the LQ and LQL models. For the evaluation of different models with different numbers of parameters we rely on the Akaike Information Criterion (AIC), a methodology widely used in radiotherapy and other fields (used for example in the study of Shuryak et al. **[30]**)

## Methods

### Clinical dataset

We have analyzed published data for patients treated for early-stage NSCLC and brain metastases.

We reviewed the data included in references [**30, 31**], disposing of some data points to increase consistency, and including some data from articles published up to November 2018. Data was discarded when Kaplan-Meier tumor control probability (TCP) values were not reported, or could not be estimated from uncorrected local tumor control rates with information from the publications, when TCP values from very different fractionation regimens were not separately specified, and when reported TCP values did not correspond to the specified time points.

In total, 116 treatment regimens were included in the database, extracting information about: 1) number of treated patients (and number of metastases for brain treatments); 2) number of fractions; 3) dose prescriptions to the isocenter and to the PTV margin, allowing estimation of the average dose to the PTV (the dose per fraction used for fitting is the average of isocenter and margin doses, as in **[30, 31]**; 4) treatment schedule; 5) Kaplan-Meier TCP values reported at 1 and 3 years after treatment for brain and lung, respectively; and 6) tumor volume when reported.

Per treatment site, the investigated regimens were:

- Lung: 61 studies (3394 patients) of early stage NSCLC, 46 of them corresponding to multifractional SBRT, 7 single-fraction SBRT, and 8 conventional fractionations.

-Brain: 55 studies (3283 patients) of brain metastasis, 30 multifractional SRS (2 of them with adjuvant whole brain radiotherapy, AWBRT), and 25 single-fraction SRS (5 of them with AWBTR).

Information about each treatment regimen is summarized in the Supplementary Materials, Tables SM1 and SM2. Further information is provided in a separated datasheet available from **[32]**.

### Radiobiological models

Our work intends to analyze if tumor control data may support the hypothesis that some death or repair mechanisms, which are not present or show little effect at the conventional doses, may arise at the high doses per fraction used in SBRT. With this purpose we performed a multimodel inference approach to compare the quality of the description of TCP data provided by alternative response models. The LQ model **[4]** was used as the reference. This model is an empirical approximation that can be derived from kinetic models describing the kinetics of DNA double-strand breaks and other lesions **[33]**. The surviving fraction of a population of cells after an acute radiotherapeutic dose *d* is,

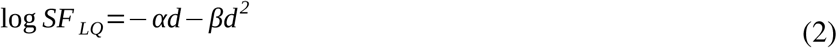

where the linear parameter, *α*, is used to model lethal damage, and the quadratic parameter, *β*, models death due to sub-lethal damage accumulation.

The validity of the LQ model is generally accepted to cover the doses per fraction ranging from 2 to 10 Gy **[5, 6]**. Beyond these limits, several modifications to the LQ model have been proposed to consider low dose hypersensitivity **[34]**, and damage saturation at high dose modifications **[10-15]**. Amongst the latter, the LQL model includes a term to compensate for the LQ underestimation of second-order repair **[11]**. For acute short irradiations, the LQL model provides the following expression for the surviving fraction, with the parameter *δ* controlling the shape of the curve at large doses:

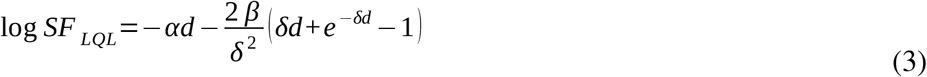

Other models have been developed to describe this relative decreasing radiosensitivity with increasing doses, like the USC model **[14]** or the Pade linear quadratic model (PLQ) **[15]**. In this work we will restrict our analysis to the LQL model among this type of models.

As mentioned above, these models aim to describe the relative decrease in effectivity in radiosensitivities with increasing doses observed in in-vitro experiments. However, there is recent experimental evidence of high doses per fraction producing higher damage than expected from low-dose extrapolation due to the activation of new mechanisms of cell death, which seems to be related to induction of vascular damage and the activation of an immune-response against the tumor **[8]**.

There have been attempts at modelling the contribution of indirect damage **[35-37]**, but there is no simple closed-form expression to model such effect. In order to study whether indirect damage contributes to observed clinical control rates, we hypothesize that an *ad hoc* modification of the LQ-model with dose-dependent alpha and beta terms, *α(d*) and *β(d*), may provide better fits to experimental dose-response curves. For example, if indirect damage is more important for large doses, *α(d*) and/or *β(d*) should have the form of monotonically-increasing functions. As there is no functional form for the functions *α(d*) and *β(d*), even though phenomenological forms could be derived from studies like **[35-37]**, we have investigated simple functional dependencies with the following form:

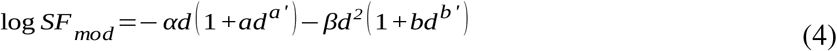

With *a’* and *b’* taking tentative values of 1/2, 1/3, 1 and 2, to explore a variety of functional dependencies. Note that single-term extensions, to the linear and quadratic terms, can be investigated by setting *b* or *a* to zero, respectively. Although the approach for this formulation is empirical, several biological processes might be associated to the proposed terms, and not only those accounting for indirect cell death: i) Negative *a* and *b* values might be related with the lack of reoxygenation occurring when tumors are treated with hypofractionaged regimens that use large doses per fraction, leading to increasing effective radioresistance with increasing dose; ii) Negative *b* values might be associated with damage saturation/repair as described by the LQL and similar models; and iii) Positive values of *a* and *b* might be in turn related with indirect tumor cell death mechanisms such as vascular damage and immune response (especially in the case of *b*, if such mechanisms are predominant at large doses). According to this reasoning, we have set *a*≤0 and no specific sign for *b* in our study.

For a treatment of ***n*** fractions (doses per fraction can be different along during the treatment), the overall surviving fraction at the end of the treatment can be written as,

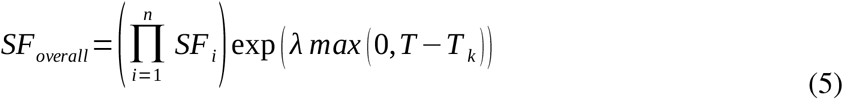

where *SF_i_* is the surviving fraction of each radiation fraction (modelled with any of the models presented above). The last term of the equation accounts for accelerated proliferation when treatment time *T* excesses the proliferation kick-off time *T_k_*.

### TCP modelling

To quantify treatment outcome, tumor control probability was modelled using a logistic function as in **[31, 38]**,

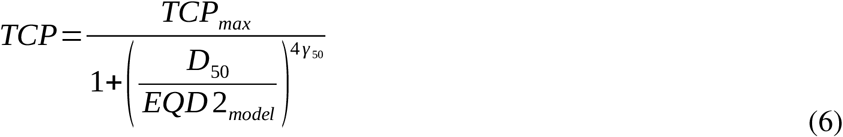

where *TCP_max_* sets an upper limit to the tumor control probability, *D*_50_ is the dose with 50% control probability and γ_50_ is the normalized dose-response gradient (that defines the setting the slope of the dose response curve). Model-derived conventional treatment (2 Gy per fraction, 5 fractions per week) equivalent doses, *EQD2_model_*, were calculated using the different models under investigation (equations 2-5) to derive issoeffective doses.

The normalized dose-response gradients were fixed to *γ*_50_=0.83 and 0.7 for NSCLC and brain metastases respectively, in accordance with previously published values calculated from clinical multi-institutional studies **[39]**. This would account for the several sources of variability affecting our database, which includes intrinsic radiosensitivity differences between tumors in different patients, intratumor heterogeneities, and differences in cohort tumor sizes and treatments between institutions.

Alternatively, we also explored the Poissonian formulation of TCP with population-averaging of radiosensitivities **[40, 41]**. For an initial number of tumor clonogens *N_0_*, the interpatient variation of intrinsic radiosensitivity in the population can be accounted for by using a probability density function for the distribution of the radiosensitivity parameter *α*:

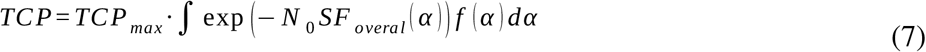

SF_overall_ is the surviving fraction given by Eq. (5). Following the approach of Shuryak et al. **[30]**, we used the Gamma distribution, which contrary to the Gaussian, converges to zero at *α*=0, as this would be more adequate to model the biological distribution of radiosensitivity in patients. The number of clonogenic cells was set to *N*_0_=10^5^, and the shape parameter of the Gamma distribution, *g*, was set to match the steepness of clinical dose-response curves (0.83 for NSCLC, 0.7 for brain).

### Evaluation of goodness of fit

Information-theoretic approaches are widely used to compare and rank multiple competing models with different number of parameters, as it is the case in this study, providing data-based strength of evidence for each of them. One example of this is the Akaike Information Criterion with sample size correction (AIC_c_), which considers the sample size and the number of model parameters to quantify the amount of Kullback-Leibler information that is lost when a model is used to approximate experimental observations **[42, 43]**:

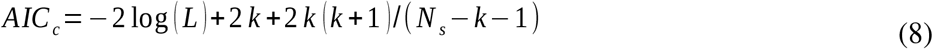

In this expression, *k* is the number of model parameters, *N_s_* is sample size (number of studies in the dose-response curve), and *L* is the likelihood function for the model:

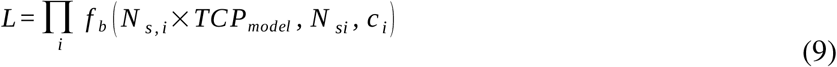

where ***i*** spans the number of points that conform the dataset, *i.e*. treatment cohorts, and *N_sj_* is the number of patients considered in each cohort. In the context of TCP modelling, the binomial probability function can be used to derive the probability, *f_b_*, that the experimental TCP value yields the number of controls estimated from the model and the number of patients involved in the study. In order to derive realistic estimates for the experimental TCP, *c*, taking into account the sample size of every cohort, binomial proportion confidence intervals were calculated from the experimental TCP data. The Agresti-Coull approximation was used for *c*, and confidence intervals were calculated with the Clopper-Pearson method by using the *p*th quantile from the beta distribution **[44, 45]**.

In this work, each radiobiological model was fitted by maximizing the likelihood over all the cohorts/fractionation regimens included in the dataset. In total, 26 models were analyzed: the LQ and LQL models, and combinations of the different functional dependencies introduced in the modified LQ model as described in equation 4.

The model exhibiting the lowest AIC_c_ is considered the best fitting model. The investigated radiobiological models were compared to the LQ model, using this latter as reference. For this, the relative likelihood of each model *j, P_j_*, given the data, was expressed in terms of the difference between the AIC_c_ value of the LQ model, taken as reference, and the AIC_c_ of the other model ΔAIC_c,j_ = AIC_c,LQ_ – AIC_c,j_:

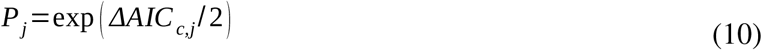

The ratio of likelihoods for any two models is the evidence ratio, *EVR*, which shows that the evidence for a model is *EVR* times stronger relative to the other. Models showing better performance than the LQ to fit TCP data would have smaller AIC_c_ values (positive ΔAIC_c_) and *EVR* >1.

A simulated annealing optimization method **[46]** was implemented in Matlab (The Mathworks, Natwick, MA) to maximize the likelihood. For the fit to the logistic model we let some parameters free *(D_50_, λ*, and *δ, a* and *b*, if used), while fixing values for some others (*α*, *α/β, T_k_, γ_50_*, and *TCP_max_*). The LQ model radiosensitivity parameter *α* was given a fixed value of 0.3 Gy^-1^, while *TCP_max_* was chosen following the approach of Jeong et al. **[31]** to replicate the experimental observation that lung and brain radiotherapy tumor control outcomes typically saturate to a value close to 95%. The *α/β* was generally fixed to 10 Gy, and *T_k_* to 28 d, within the range of reported values **[31, 47, 48]**, but other values were also explored. The Poisson TCP expression was also explored to confirm the results achieved for the different models under the logistic TCP formulation. In this case, the radiosensitivity parameter *α* was optimized instead of *D_50_*. Therefore, both TCP models have the same number of free parameters.

A more detailed presentation of the fitting procedure and the statistical model evaluation is included as part of the Supplementary Materials.

### Stability of results under variation of parameters and dose/response perturbations

The quality of the fits corresponding to the different models can be affected by the values assigned to the parameters that are fixed in the optimization process. To investigate this, several values were tested for some parameters, namely: i) the *α/β* ratio; ii) the kick-off time for accelerated repopulation, *T_k_* and; iii) the parameters setting the slope of the TCP curve, *γ_50_* and *g*, for Logit and Poisson respectively. The parameter values tested in these simulations are summarized in Table SM3.

We also studied the sensitivity of the fits to uncertainties in our database (dose and TCP values), the aim being to explore whether the rank/performance of models may be affected by inherent experimental uncertainties (i.e. the robustness of the results). Dose and TCP values were perturbed, either alone or in combination, and the optimization process and AIC_c_ analysis performed with the perturbed dataset. This procedure was repeated 20 times (50 times when dose and TCP values where simultaneously perturbed). For dose perturbations, we chose a normal distribution with a mean equal to the dose per fraction and a 5% relative standard deviation. Tumor control probability values were sampled from the binomial distribution, using the Agresti-Coull probability associated to each cohort and the number of patients involved.

The effects of dose/TCP perturbations to the database were additionally analyzed with hypothesis testing. In this case, we tested if the proportion of instances that a model showed better performance than the LQ was significant, i.e. better than chance with a 0.5 probability, by using the one-proportion z-test at a 95% confidence level.

### Implementation

Different routines for fitting and analyzing the results were implemented in Matlab. The code and data in Matlab format, and the experimental dataset (in openoffice table format) are available from the Harvard Dataverse repository **[32]**.

## Results

### Dose-response curve of NSCLC shows evidence of increasing radiosensitivity with increasing dose

The study of NSCLC dose response data showed improved fit quality for the modified LQ model, compared with the LQ. This trend was observed both with the logistic and Poisson TCP formulations. The modified LQ model with b’=1/2 is the one that provides the best results, with b>0 (*b* =0.10 and *b* =0.06 for logit and Poisson fits, respectively, see Table SM4), which implies an increasing *β(d*) with increasing dose. Consistently with this, the LQL model, which describes a decreasing effectivity with increasing dose, was not able to improve the quality of the fit achieved with the LQ model. In Figure 1 we present best-fits to TCP data with the LQ and modified LQ models, and in Table 1 we report AIC_c_ and EVR values for a selection of models.

**Figure 1:**
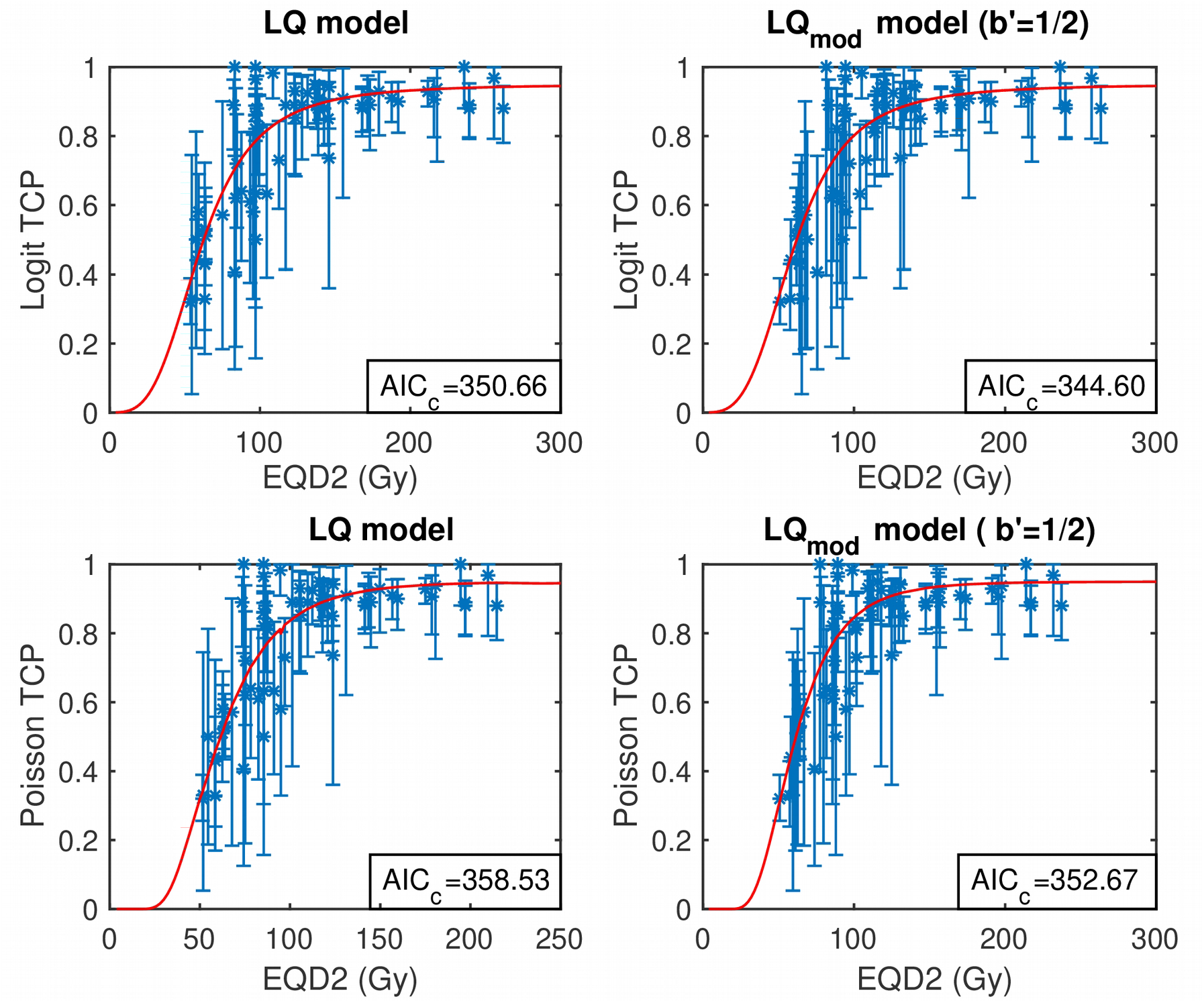
Best fits to early-stage NSCLC dose response data under logistic (top panels) and Poisson TPC formulation (bottom panels) using the LQ model (left) and the LQ modification with functional dependency b’=1/2 (right).

**Table 1:** Akaike-information-criterium (AIC_c_) and evidence ratio (EVR) values for the fits of the logit/Poisson TCP models (based on the LQ model, taken as reference, the LQL model, and modifications of the LQ models) to the early stage NSCLC and brain metastases datasets.

|  | NSCLC |  |  |  | Brain Metastases |  |  |  |
| --- | --- | --- | --- | --- | --- | --- | --- | --- |
|  | Logit model |  | Poisson model |  | Logit model |  | Poisson model |  |
|  | AIC<sub>c</sub> | EVR | AIC<sub>c</sub> | EVR | AIC<sub>c</sub> | EVR | AIC<sub>c</sub> | EVR |
| **LQ** | 350.66 | 1.00 | 358.53 | 1.00 | 825.61 | 1.00 | 844.75 | 1.00 |
| **LQL** | 352.76 | 0.35 | 359.72 | 0.55 | 701.62 | 8.4E+26 | 709.63 | 2.2E+29 |
| **LQ ext (a'=1/2)** | 352.87 | 0.33 | 360.41 | 0.39 | 827.85 | 0.33 | 846.45 | 0.43 |
| **LQ ext (a'=1)** | 352.88 | 0.33 | 360.24 | 0.43 | 788.43 | 1.2E+08 | 804.11 | 6.7E+08 |
| **LQ ext (b'=1/2)** | 344.60 | 20.68 | 352.67 | 18.70 | 702.46 | 5.5E+26 | 711.91 | 7.0E+28 |
| **LQ ext (b'=1)** | 348.65 | 2.72 | 356.69 | 2.51 | 703.68 | 3.4E+10 | 711.38 | 9.1E+28 |
| **LQ ext (a'=1/2, b'=1/2)** | 346.91 | 6.52 | 354.98 | 5.88 | 704.75 | 1.7E+26 | 713.17 | 3.7E+28 |
| **LQ ext (a'=1/2, b'=1)** | 350.92 | 0.88 | 358.77 | 0.89 | 705.98 | 9.5E+25 | 713.41 | 3.3E+28 |
| **LQ ext (a'=1, b'=1/2)** | 346.86 | 6.68 | 354.95 | 5.98 | 704.75 | 1.7E+26 | 712.78 | 4.5E+28 |
| **LQ ext (a'=1, b'=1)** | 350.95 | 0.87 | 358.70 | 0.92 | 706.04 | 9.2E+25 | 713.33 | 3.4E+28 |

Although the fitting improvement cannot be easily noticed by visual inspection (see Figure 1), the AIC_c_ supports the difference in fit quality, with ΔAIC_c_~6 and EVR~20 for the modified LQ model with b’=1/2.

Further detailed information is provided in the Supplementary Materials: in Figures SM1 and SM2 we present best fits (TCP vs EQD_2_) for each model, for logit and Poisson descriptions of the TCP; best-fitting parameters for each model in Table SM4, and; AICs and EVR values for each model in Table SM5.

### Dose-response curve of brain metastases shows evidence of damage saturation with increasing dose

Analysis of brain metastases data shows that the LQL provides the best fits, particularly compared with the LQ. The results obtained with the LQL model are very similar to those obtained with the modified LQ model with b’=1/2 and b<0 (b= −0.13 for logit and Poisson, Table SM6), which is not unexpected as this model behaves similarly to the LQL model. In Figure 2 we present best-fits for those models, with the Poisson and logit TCP methodologies.

**Figure 2:**
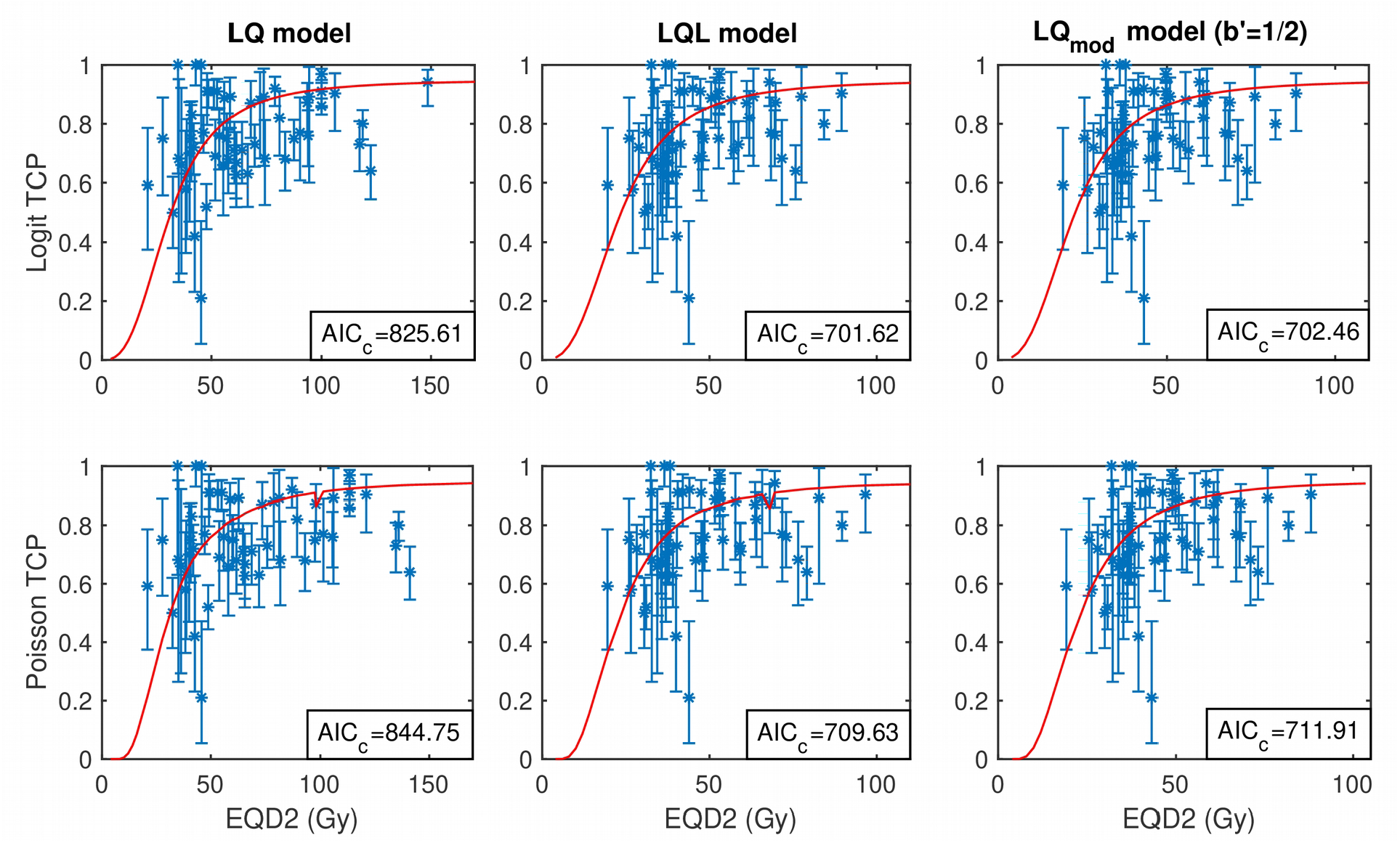
Best fits to brain metastases dose response data under logistic (top panels) and Poisson (bottom panels) TPC formulation using the LQ model (left), the LQL model (center) and the LQ modification with functional dependency b’=1/2 (right).

While the fits improvement is again hardly noticeable in the figures, it can be easily observed in Table 1, which reports AIC_c_ and EVR values for a selection of models. In this case, ΔAIC_c_ of the best model (the LQL) are far larger than in the case of NSCLC, ΔAIC_c_~125, which results in values of EVR>10^20^.

As for NSCLC, we report detailed information of the results in the Supplementary Materials: in Figures SM3 and SM4 we present best fits (TCP vs EQD_2_) for each model, for logit and Poisson descriptions of the TCP; AICs and EVR values for each model in Table SM5, and; best-fitting parameters for each model in Table SM6.

### Stability of results under variation of parameters

In Table 2 we present AIC_c_ and EVR values for the best-fitting models (modified LQ model with b’=1/2 for NSCLC and brain, and LQL for brain) and the LQ model, when different fixed values are given to parameters *T_k_, α/β* and γ*_50_*. Results reported in this table correspond to the logistic TCP methodology.

**Table 2:** Effect of variations of fitting parameters *T_k_, α/β* and *γ* on the evidence ratio (EVR) of the modified LQ model with b’=1/2 (NSCLC and brain metastases) and the LQL (brain metastases), relative to the LQ model. AIC_c_ values for the LQ model fit are also reported, to show fit improvements with parameter variations. Parameters that were not varied were set to *T_k_* = 28 d, *α/β* = 10 Gy and *γ*_50_=0.83.

|  | NSCLC |  |  | Brain Metastases |  |  |  |  |
| --- | --- | --- | --- | --- | --- | --- | --- | --- |
|  | AIC<sub>c</sub><br>LQ | AIC<sub>c</sub> LQmod<br>b'=1/2 | EVR (LQmod<br>b'=1/2) | AIC<sub>c</sub><br>LQ | AIC<sub>c</sub><br>LQL | AIC<sub>c</sub> LQmod<br>b'=1/2 | EVR<br>(LQL) | EVR (LQmod<br>b'=1/2) |
| **$T_k$ variation** |  |  |  |  |  |  |  |  |
| $T_k = 28$ d | 350.7 | 344.60 | 20.68 | 825.6 | 701.62 | 702.46 | 8E+26 | 6E+26 |
| $T_k = 21$ d | 348.4 | 344.40 | 7.33 | 825.6 | 701.66 | 702.45 | 8E+26 | 6E+26 |
| $T_k = 14$ d | 345.4 | 343.56 | 2.56 | 825.6 | 701.67 | 702.43 | 8E+26 | 6E+26 |
| $T_k = 7$ d | 345.3 | 343.90 | 1.98 | 825 | 679.68 | 676.21 | 4E+31 | 2E+32 |
| **$\alpha/\beta$ variation** |  |  |  |  |  |  |  |  |
| $\alpha/\beta = 20$ Gy | 420.3 | 350.11 | 2E+15 | 752.2 | 709.45 | 711.55 | 2E+09 | 7E+08 |
| $\alpha/\beta = 10$ Gy | 350.7 | 344.60 | 20.68 | 825.6 | 701.62 | 702.46 | 8E+26 | 6E+26 |
| $\alpha/\beta = 5$ Gy | 339.7 | 335.32 | 8.95 | 917.1 | 694.53 | 696.62 | 2E+48 | 8E+47 |
| **$\gamma_{50}$ variation** |  |  |  |  |  |  |  |  |
| $\gamma_{50} = 1.5$ | 369.7 | 371.89 | 0.34 | - | - | - | - | - |
| $\gamma_{50} = 1$ | 347.5 | 346.55 | 1.60 | 963.6 | 776.16 | 766.28 | 5E+40 | 7E+42 |
| $\gamma_{50} = 0.83$ | 350.7 | 344.60 | 20.68 | 888.4 | 732.90 | 731.83 | 6E+33 | 1E+34 |
| $\gamma_{50} = 0.7$ | 355.2 | 344.92 | 170.85 | 825.6 | 701.62 | 702.46 | 8E+26 | 6E+26 |
| $\gamma_{50} = 0.3$ | - | - | - | 658.3 | 646.99 | 647.33 | 289 | 2E+02 |

In NSCLC, the faster the start of accelerated proliferation (the lower value of *T_k_*), the lower the evidence ratio of the modified LQ model. Proliferation can be viewed as an increasing relative radiosensitivity with increasing dose, as treatments with low doses per fraction and long treatment times will lose effectiveness due to proliferation, while treatments with high doses per fraction and short treatment times will not. This is the same behavior modeled by the modified LQ model with b’=1/2 and b<0. Therefore, increasing the contribution of proliferation causes the superiority of the modified LQ model over the LQ model to fade away. ΔAIC_c_ values decrease from 6 (EVR~21) for *T_k_=28* d to 1.4 (EVR~2) for T_k_=7 d. The same trend is observed with decreasing values of *α/β.* ΔAIC_c_ values decrease from 70 (EVR~2×10^8^) for *α/β=*20 d to 4.4 (EVR~9) for *α/β=5* Gy. A different trend is observed when changing the value of γ_50_, which models the slope of the dose-response curve: the superiority of the modified LQ model over the LQ model diminished with increasing *γ*_50_ (steeper slopes), and totally fades away when *γ*_50_=1.5.

On the other hand, for brain metastases the superiority (ΔAIC_c_ and EVR values) of the LQL over the LQ model is not importantly affected by variations of *T_k_, α/β* and *γ* with values of ΔAIC_c_> 11 and EVR>289 for every investigated set of parameters.

As mentioned above, these results correspond to fits performed with the logit TCP methodology, but identical trends are observed when using the Poisson methodology, and shown in Tables SM7 and SM8.

### Stability of results under dose/response perturbations

The quality of the fits changes drastically when dose/TCP values are perturbed, as can be noticed in the mean and standard deviation of AIC_c_ values for the LQ model reported in Table 3. Accordingly, EVR and ΔAIC_c_ values suffer large variations, but the modified LQ model (for NSCLC) and LQL (for brain) remain systematically better than the LQ model in spite of dose/TCP perturbation.

In the case of NSCLC, the modified LQ model is superior to the LQ model in 20/20 experiments with dose perturbations, 17/20 with TCP perturbations, and 33/50 experiments with both perturbations. The z-test of proportions shows that the superiority of the former model is significant, wit *p*<10^-5^, *p*<10^-3^, and *p*=0.01, respectively. In the case of brain, dose/TCP perturbations do not affect at all the rank of the LQL model, which is superior in 20/20 (p<10^-5^), 20/20 (p<10^-5^), and 50/50 (p<10^4^°) experiments with dose, TCP, and dose and TCP perturbations, respectively.

**Table 3:** Mean ± one standard deviation of AIC_c_, ΔAIC_c_ and EVR values for fits with the LQ model, LQL model (for brain metastases) and modified LQ model with b’=1/2 (for NSCLC) when doses, tumor control probabilities, or both values in the dataset were perturbed. Fits were performed with the logistic TCP formulation using *T_k_* = 28, *α/β*=10 Gy and *γ*_50_=0.83 (NSCLC) or 0.7 (brain).

|  | NSCLC |  |  | Brain Metastases |  |  |
| --- | --- | --- | --- | --- | --- | --- |
| | AIC<sub>c</sub> LQ | ΔAIC<sub>c</sub><br>LQ<sub>mod</sub><br>b'=1/2 | EVR (LQ<sub>mod</sub><br>b'=1/2) | AIC<sub>c</sub> LQ | ΔAIC<sub>c</sub> LQL | EVR (LQL) |
| **Dose Perturbations** | 375 ± 17 | 11.4 ± 6.5 | 8E+05 ± 3E+06 | 818 ± 31 | 119.5 ± 18.1 | 2E+31 ± 8E+31 |
| **TCP perturbations** | 384 ± 21 | 2.2 ± 3.1 | 36 ± 147 | 832 ± 50 | 114.2 ± 23.7 | 1E+35 ± 5E+35 |
| **Dose & TCP perturbations** | 409 ± 29 | 4.2 ± 6.6 | 2479 ± 9163 | 849 ± 51 | 119.6 ± 30.4 | 2E+39 ± 1E+40 |

## Discussion

Pre-clinical experiments have shown that vascular damage and immune effects trigger indirect tumor cell death. This may play a role in high-dose SBRT/SRS protocols that deliver few fractions per treatment, or even only one. In addition, saturation of damage (due to damage repair) may also play a role at large doses. In this work we have analyzed a large dataset of dose-response for NSCLC and lung metastases, looking for evidence of *indirect* damage and/or damage saturation in the experimental data. The methodology that we follow consists on fitting dose-response curves to response models based on the conventional LQ model, the LQL model (with decreasing relative radio-sensitivities with increasing dose) and on simple modifications of the LQ model to allow for increasing relative radio-sensitivities with increasing dose. If mechanisms accounting for decreasing radio-sensitivity with increasing dose are dominant (like damage saturation or lack of reoxygenation) the LQL should prove superior; but if contribution of indirect damage with increasing dose is dominant, modifications of the LQ should prove superior; and if none of those contributions is important, the LQ should be the best model.

For the NSCLC dataset, a modification of the LQ model with a slow increase of the beta term with dose (~ √***d*)** provides the best fit to experimental data (ΔAIC_c_~6, EVR~20):

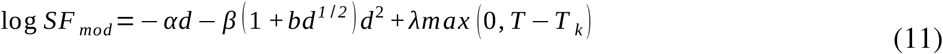

This could be interpreted as a signal of contribution of indirect cell damage at large doses per fraction, as observed in experimental studies **[8, 16, 17]**. On the other hand, fitting the dataset of brain metastases showed the LQL model to be the best, which points out a relative decrease of radiosensitivity with increasing dose.

There are several confounding factors that could affect these conclusions and need to be studied. For example, if accelerated proliferation kicks off fast, it would create a net effect similar to that observed in the NSCLC dataset - treatments with few fractions and large doses per fraction will not be affected (or lightly affected) by proliferation, an effect that can be interpreted as an increasing relative radiosensitivity with increasing dose. In fact, we have observed that by decreasing the proliferation kick-off time from the typical 4 weeks values, the statistical superiority of the modified LQ-model also decreases. However, we need to reach values of ***T****k* of 1 week in order to eliminate the superiority of the modified LQ model (ΔAIC_c_~1.4, EVR~2). Such fast kick-off times do not seem compatible with reported times for NSCLC **[31, 47]**.

Different *α/β* values can also affect the fits. We have investigated values of *α/β* =10 (primarily), 5 and 20 Gy, a large range where the *α/β* of this tumors should lie **[48]**. Interestingly, increasing the *α/β* ratio from 10 Gy to 20 Gy does not improve the quality of the fits (AIC_c_=345 for 10 Gy, AIC_c_=350 for 20 Gy for the modified LQ model in NSCLC, AIC_c_=702 for 10 Gy, AIC_c_=712 for 20 Gy for the LQL model in brain metastases). The ΔAIC_c_ values of the best models tend to diminish with decreasing *α/β*, values, yet we should get to values below 5 Gy for the differences to vanish, and those values seem not to be experimentally justified.

We have also evaluated whether differences in volumes of tumors may introduce a systematic bias in our analysis by investigating volumes reported in the experimental studies. If small NSCLC tumors are prescribed larger doses per fraction than large tumors, it could be interpreted as an increasing relative radiosensitivity with increasing doses, because small tumors are easier to control. The opposite should happen in brain metastases in order to explain the superiority of the LQL model. We have analyzed reported volumes (mean/median values) in the experimental studies, finding no association between volume and prescribed dose per fraction: nor a Spearman test of volume-dose data neither a hypothesis-test of tumors below ***V_t_*** receiving higher doses showed significance (threshold volumes ***V****_t_* were set to the median of the tumor volumes amongst all the cohorts, 25 cm^3^ and 2 cm^3^ for lung and brain, respectively). These results are shown in Figure SM5.

Saturation of sub-lethal repair mechanisms at high doses might also result in an increasing effective beta-term with increasing dose as observed in NSCLC. However, this effect is not expected to contribute at these dose values **[29]**.

Experimental TCP values are subject to statistical uncertainties due to the finite size of the patient pool, and uncertainties in the doses may be large, especially because dose distributions in the tumors are highly heterogeneous while we report a scalar dose value (mean of edge and isocenter doses). We have investigated the effect of dose/TCP uncertainties in the fitting procedure, finding that the best models (modified LQ and LQL for NSCLC and brain metastases, respectively) remain the best under TCP/dose perturbations. A z-test of proportions showed that the superiority of those models over the LQ model is significant.

A recent study **[30]** found that the LQ-Poisson model provides a better fit to dose-response curves for NSCLC and brain metastases than the LQL model (and other models like the PLQ and USC). Our results are in disagreement with this study for brain metastases, where we have observed that the LQL is superior to the LQ. The origin of this disagreement might be on the different datasets used in both studies. However, we also attribute part of the disagreement to the management of averaging of radiosensitivities, as Shuryak et al only included what is called intra-tumor averaging in the computation of Poisson-TCPs, which cannot bend the slope of the response curve as the interpatient does. This results in very steep dose-response curves for the LQL/USC/PLQ models, and very large AIC_c_ values for those models, and therefore large negative ΔAIC_c_ values when compared to the LQ model. On the other hand, all models tested in reference **[30]** are models with decreasing radio-sensitivities with increasing dose. Therefore, the authors could not detect the effect of increasing radio-sensitivities with increasing dose in early stage NSCLC that we have found in this work.

Our results should be approached with care, for there are confounding factors that can affect our conclusions. Several of such factors, including proliferation, dose/TCP uncertainties and a systematic volume effect, have been investigated and do not seem to compromise the conclusions of our study. The significance of the differences among models can also be questioned. When using the Akaike methodology, models with ΔAIC_c_ values below 2 (*EVR*<2.7) are generally considered to be as good as the reference model, while between 2-6 (EVR<20) models are rarely dismissed as they would still have some support. Although there is still some debate about when a model can be considered implausible, model rejection is usually considered when ΔAIC_c_ is above 6 (*EVR*>20), or 9-11 ***(EVR*** 90-245) in more conservative approaches **[49, 50, 30]**. The modified LQ model for NSCLC has ΔAIC_c_ ~ 6, which is borderline significant according to the above definitions. However, the LQL model (and the modified LQ with b’=1/2 and negative ***b*** values) for brain metastases has ΔAIC_c_ ~125, a result that should be considered significant. However, it is important to point out that fits to brain data are significantly worse than fits to NSCLC data: AIC_c_ ~ 700 vs. 350. This is probably caused by the larger heterogeneity of brain metastases, for example originating from different types of primary tumors. It should not be discarded that such heterogeneities might be affecting the significant superiority of the LQL model.

In this work, we have found evidence that a modification of the LQ model with a beta-term increasing with dose provides better fits to NSCLC dose-response curves than the LQ and LQL models. This might be interpreted as an indirect cell damage contribution in the response of NSCLC. On the other hand, the analysis of the dose-response curve of brain metastases did not show the same behavior. In the latter case, the LQL model provided the best first, which points out a saturation of damage with increasing dose.

Our results should trigger further research in this area, aiming at identifying and characterizing the effect of indirect cell damage in clinical tumor response, and at developing novel biomathematical models to describe such effects. These results can assist on the design of optimal radiotherapy treatments for NSCLC and brain metastases, aiming at avoiding over- or under-treatment. Dose prescription to such tumors may be re-evaluated according to the reported evidence.

## Data Availability

The code and data are available from the Harvard Dataverse repository.

https://dataverse.harvard.edu/dataset.xhtml?persistentId=doi:10.7910/DVN/RRIJHF

**List of abbreviations**
NSCLC: Non Small Cell Lung Cancer
LQ: linear quadratic
LQL: lineal quadratic lineal
AIC: Akaike Information Criterion
TCP: Tumor Control Probability
SRS: Stereotactic RadioSurgery
SBRT: Stereotactic Body Radiation Therapy
AWBRT: adjuvant whole brain radiotherapy
PLQ: Pade linear quadratic

## Declarations

### Ethics approval and consent to participate

Not applicable

### Consent for publication

Not applicable

### Availability of data and materials

The datasets generated and/or analysed during the current study are available in the Harvard Dataverse repository, doi.org/10.7910/DVN/JKMHMR

### Authors’ contributions

AGA wrote the code, collected, analyzed and interpreted data, and drafted the manuscript. SN contributed to code implementation and data analysis. MP contributed to data collection, analysis and interpretation. AGC contributed to data collection, analysis and interpretation. JPM developed the study concept, supervised the project, analyzed and interpreted data, and drafted the manuscript. All authors revised and approved the final version of the manuscript.

## References

1. Sperduto PW. A review of stereotactic radiosurgery in the management of brain metastases. Technol Cancer Res Treat 2003;2:105–110.

2. Chang BK. Timmerman RD. Stereotactic body radiation therapy: a comprehensive review. Am J Clin Oncol 2007;30:637–44.

3 Lo SS. Fakiris AJ. Chang EL. Mayr NA. Wang JZ. Papiez L. et al. Stereotactic body radiation therapy: a novel treatment modality. Nat Rev Clin Oncol 2010;7:44–54.

4. Fowler JF. The linear-quadratic formula and progress in fractionated radiotherapy. Br J Radiol 1989;62:679–694

5. Kirkpatrick JP. Meyer JJ. Marks LB. The linear-quadratic model is inappropiate to model high dose per fraction effects in radiosurgery. Semin Radiat Oncol 2008:18;240–3.

6. Brenner DJ. The linear-quadratic model is an appropriate methodology for determining isoeffective doses at large doses per fraction. Semin Radiat Oncol 2008:18;234–9.

7. Song CW. Park H. Griffin RJ. Levitt SH. Radiobiology of Stereotactic Radiosurgery and Stereotactic Body Radiation Therapy. In: Lewitt SH. et al. Technical basis of Radiation Therapy: Practical Clinical Application. New York: Springer Publishing Co; 2012. p. 51–61.

8. Sperduto PW. Song CW. Kirkpatrick JP. Glatstein E. A hypothesis: indirect cell death in the radiosurgery era. Int J Radiat Oncol Biol Phys 2015:91;11–13.

9. Song CW. Kim M. Cho LC. Dusenbery K. Sperduto PW. Radiobiological basis of SBRT and SRS. Int J Clin Oncol 2014:19;570–8

10. Guerrero M. Carlone M. Mechanistic formulation of a lineal-quadratic-linear (LQL) model: split-dose experiments and exponentially decaying sources. Med Phys 2010;37:4173–4181.

11. Guerrero M. Li XA. Extending the linear-quadratic model for large fraction doses pertinent to stereotactic radiotherapy. Phys Med Biol 2004;49:4825–4835.

12. Carlone MC. Wilkins D. Raaphorst GP. The modified linear-quadratic model of Guerrero and Li can be derived from amechanistic basis and exhibits linear-quadratic-linear behaviour. Phys Med Biol 2005;50:L9–L13.

13. Wang JZ. Huang Z. Lo SS. Yuh WTC. Mayr NA. A generalized linear-quadratic model for radiosurgery, stereotactic body radiation therapy, and high-dose rate brachytherapy. Sci Transl Med 2010; 2:1–7.

14. Park C. Papiez L. Zhang S. Story M. Timmerman RD. Universal survival curve and single fraction equivalent dose: useful tools in understanding potency of ablative radiotherapy. Int J Radiat Oncol Biol Phys 2008;70:847–52.

15. Belkic D. Parametric analysis of time signals and spectra from perspectives of quantum physics and chemistry. Adv Quant Chem 2011;61:145–260.

16. Song CW. Lee YJ. Griffin RJ. Park I. Koonce NA. Hui S. et al. Indirect Tumor Cell Death After HighDose Hypofractionated Irradiation: Implications for Stereotactic Body Radiation Therapy and Stereotactic Radiation Surgery. Int J Radiat Oncol Biol Phys 2015;93:166–72.

17. García-Barros M. Paris F. Cordón-Cardo C. Lyden D. Rafii S. Haimovitz-Friedman A. et al. Tumor response to radiotherapy regulated by endothelial cell apoptosis. Science 2003;300:1155–9.

18. Park HJ. Griffin RJ. Hui S. Levitt SH. Song CW. Radiation-induced vascular damage in tumors: implications of vascular damage in ablative hypofractionated radiotherapy (SBRT and SRS). Radiat Res 2012;177:311–27.

19. Song CW. Glatstein E. Marks LB. Emami B. Grimm J. Sperduto PW. et al. Biological Principles of Stereotactic Body Radiation Therapy (SBRT) and Stereotactic Radiation Surgery (SRS): Indirect Cell Death. Int J Radiat Oncol Biol Phys. 2019 IN PRESS

20. Lee Y. Auh SL. Wang Y. Burnette B. Wang Y. Meng Y. et al. Therapeutic effects of ablative radiation on local tumor require CD8+ T cells: changing strategies for cancer treatment. Blood 2009:114;589–95.

21. Filatenkov A. Baker J. Mueller A. Kenkel JA. Ahn G-O. Dutt S. et al. Ablative tumor radiation can change the tumor immune cell microenvironment to induce durable complete remissions. Clin Cancer Res 2015;21(16):3727–39.

22. De La Maza L. Wu M. Wu L. Yun H. Zhao Y. Cattral M. et al. *In Situ* Vaccination after Accelerated Hypofractionated Radiation and Surgery in a Mesothelioma Mouse Model. Clin Cancer Res. 2017;23(18):5502–5513.

23. Wang Y. Liu Z. Yuan H. Deng W. Li J. Huang Y. et al. The reciprocity between radiotherapy and cancer immunotherapy. Clin Cancer Res 2019;25(6):1709–1717

24. Herrera FG. Bourhis J. Coukos G. Radiotherapy combination opportunities leveraging immunity for the next oncology practice. CA Cancer J Clin. 2017;67(1):65–85.

25. He J. Yin Y. Luster TA. Watkins L. Thorpe PE. Antiphosphatidylserine antibody combined with irradiation damages tumor blood vessels and induces tumor immunity in a rat model of glioblastoma. Clin Cancer Res. 2009;15(22):6871–80.

26. Deng L. Liang H. Burnette B. Beckett M. Darga T. RR Weichselbaum, Fu Y-X. Irradiation and anti-PD-L1 treatment synergistically promote antitumor immunity in mice. J Clin Invest 2014;124(2):687–95.

27. Moding EJ. Castle KD. Perez BA. Oh P. Min HD. Norris H. et al. Tumor cells, but not endothelial cells, mediate eradication of primary sarcomas by stereotactic body radiation therapy. Sci Transl Med 2015;7:278ra34.

28. Torok JA. Oh P. Castle KD. Reinsvold M. Ma Y. Luo L. et al. Deletion of ATM in tumor but not endothelial cells improves radiation response in a primary mouse model of lung adenocarcinoma. Cancer Res. 2019;79:773–782

29. Brown JM. Carlson DJ. Brenner DJ. The tumor radiobiology of SRS and SBRT: are more than the 5 Rs involved?. Int J Radiat Oncol Biol Phys 2014;88:254–62.

30. Shuryak I. Carlson DJ. Brown M. Brenner DJ. High-dose and fractionation effects in stereotactic radiotherapy: Analysis of tumor control data from 2965 patients. Radiother Oncol 2015;115:327–334.

31. Jeong J. Oh JH. Sonke JJ. Belderbos J. Bradley JD. Fontanella AN. et al. Modeling the Cellular Response of Lung Cancer to Radiation Therapy for a Broad Range of Fractionation Schedules. Clin Cancer Res 2017;23:5469–79.

32. Gago-Arias A. Neira S. Pombar M. Gómez-Caamaño A. Pardo-Montero J. Replication data for “Evaluation of indirect damage and damage saturation effects in dose-response curves of hypofractionated radiotherapy of early-stage NSCLC and brain metastases”. Harvard Dataverse 2020. doi.org/10.7910/RRIJHF

33. Curtis SB. Lethal and potentially lethal lesions induced by radiation-a unified repair model Radiat. Res. 1986;106:252–270.

34. Joiner MC. Marples B. Lambin P. Short SC. Turesson I. Low-dose hypersensitivity: current status and possible mechanisms. Int J Radiat Oncol Biol Phys 1997;49:379–389.

35. Serre R. Benzekry S. Padovani L. Meille C. André N, Ciccolini J. et al. Mathematical Modeling of Cancer Immunotherapy and Its Synergy with Radiotherapy. Cancer Res. 2016;76(17):4931–40.

36. Paul-Gilloteaux P. Potiron V. Delpon G. Supiot S. Chiavassa S. Paris F. et al. Optimizing radiotherapy protocols using computer automata to model tumour cell death as a function of oxygen diffusion processes. Sci Rep 2017;7(1):2280.

37. Rodríguez-Barbeito P. Díaz-Botana P. Gago-Arias A. Feijoo M. Neira S. Guiu-Souto J. et al. A model of indirect cell death caused by tumor vascular damage after high-dose radiotherapy. Cancer Research 2019;79(23): 6044–53.

38. Bentzen SM. Tucker SL. Quantifying the position and steepness of radiation dose-response curves. Int J Radiat Biol 1997;71:531–542

39. Okunieff P. Morgan D. Niemierko A. and Suit HD. Radiation dose-response of human tumors. Int J Radiat Oncol Biol Phys 1995;32(4):1227–1237.

40. Webb S. Nahum AE. A model for calculating tumour control probability in radiotherapy including the effects of inhomogeneous distributions of dose and clonogenic cell density. Phys Med Biol. 1993;38(6):653–66.

41. Nahum AE. Sanchez-Nieto B. Tumor control probability modeling: Basic principles and applications in treatment planning. Physica Medica 2001;17:13–23.

42. Akaike H. A new look at the statistical model identification. IEEE Transactions on Automatic Control. 1974;19(6): 716-723.

43. Gordon RA. Regression Analysis for the Social Sciences, Routledge 2015.

44. Vollset SE. Confidence intervals for a binomial proportion. Statistics in medicine, 1993;12(9):809–824.

45. Agresti A. Coull BA. Approximate is Better than “Exact” for Interval Estimation of Binomial Proportions. Am Stat 1998;52:119–126.

46. Kirkpatrick S. Gelatt CD. Vecchi MP. Optimization by Simulated Annealing. Science. 1983;220:671–680

47. Fenwick JD. Nahum AE. Malik ZI. Eswar CV. Hatton MQ. Laurence VM. et al. Escalation and intensification of radiotherapy for stage III non-small cell lung cancer: opportunities for treatment improvement. Clin Oncol (R Coll Radiol). 2009;21(4):343–60.

48. Klement RJ. Sonke JJ. Allgäuer M. Andratschke N. Appold S. Belderbos J. et al. Estimation of the a/ß ratio of non-small cell lung cancer treated with stereotactic body radiotherapy. Radiother Oncol. 2020;142:210–216.

49. Symonds MR. Moussalli A. A brief guide to model selection, multimodel inference and model averaging in behavioural ecology using Akaike’s information criterion. Behavioral Ecology and Sociobiology 2011;65(1):13–21.

50. Burnham KP. Anderson DR. and Huyvaert KP. AIC model selection and multimodel inference in behavioral ecology: some background, observations, and comparisons. Behavioral ecology and sociobiology 2001;65(1):23–35.

